# Megapractices: an update and tentative typology of the new models of primary care provision in Scotland

**DOI:** 10.64898/2026.02.09.26345886

**Authors:** Heather McAdam, Benjamin Hunter, David Blane, Rebecca Riddell, Christopher Johnstone, Gerry McCartney

## Abstract

**Background:** GP practices in Scotland are changing with the emergence of megapractices. We aimed to update analyses of GP practice sizes in Scotland, and to begin the development of a typology of GP practices.

**Methods:** Four methods were employed: 1. Analyses of routinely published data on GP practice sizes and listed GPs to identify and quantify megapractices; 2. Qualitative interviews; 3. Creation of commercial profiles; 4. Derivation of a GP practice typology.

**Results:** Most Scottish practices have less than 9,000 patients, but five megapractices with over 30,000 patients were identified. One had grown by over 18,000 patients in two years to almost 120,000 patients, with the next largest at almost 73,000 patients. Megapractices did not engage with interviews, but commercial profiling revealed an array of complex legal and financial arrangements for several of them. We suggest that the key parameters for describing practices should focus on: ownership, legal status and motives; the extent of service provision by doctors or Allied Health Professionals (AHPs); and practice list size. We tentatively propose four common practice types in Scotland: traditional practices, megapractices, social enterprise practices, and direct NHS provision, recognising substantial diversity amongst megapractices.

**Implications:** Policymakers should carefully consider the risks of the current drift in approach to GP provisioning. Evaluation of the impacts of new service models on healthcare access, health outcomes and inequalities is urgently needed.

## Background

In Scotland, new models of provisioning General Practice (GP) services are emerging. Until relatively recently, almost all GP practices in Scotland followed a traditional model of providing services in a tightly defined geographical area, for up to c.15,000 patients, with up to c.10 GP partners employing a range of staff (including nurses, receptionists and a practice manager), working for NHS Boards (and more recently Health and Social Care Partnerships – entities created with joint accountability to NHS Boards and Local Authorities) as independent contractors. Without any policy initiative promoting change,^1^ ‘megapractices’ have emerged over the last 10 years with much larger patient lists and a quite different service model.^2^ Despite this, we know very little about the characteristics of megapractices, whether there is variation amongst megapractices in terms of their service models, or if this is having any impacts on service access, health outcomes or changes in inequalities (either in access or outcomes).^2^

It is recognised that there was a GP staffing crisis in Scotland from the late-2010s.^3^ There were increasingly frequent reports of difficulties replacing retiring GP partners, and this led to many practices being dissolved and contracts being handed back to Boards.^4^ Some Boards responded by directly employing GPs and running practices as ‘2C practices’ (in reference to the section of the 1978 NHS (Scotland) Act governing this process), but there seemed to be little appetite for this to become widespread as this was perceived to be a more expensive option for Boards. Instead, a small number of practices changed their practice model to become ‘GP-light’ by employing a large number of Allied Health Professionals (AHPs, including paramedics and nurses) and started to bid for these vacant contracts.^3^ As a consequence, megapractices emerged with rapidly increasing patient list sizes, with one reaching a list size of 77,000, and another 101,000, by 2021.^2^ This trend has been accompanied by growing dissatisfaction expressed by the public about difficulties in getting appointments to see GPs and difficulties in finding GPs who will take on new patients in some areas.^5^

In this contribution, we aimed to update the analyses of GP practice sizes in Scotland, and to begin the development of a typology of GP practices to reflect this evolving trend towards large practices and new practice models.

## Methods

We employed four different methods in this work, each of which is detailed separately below.

### Update of trends in the size of GP practices in Scotland

Data on practice list sizes in Scotland are published by Public Health Scotland. This provides a breakdown of registered patients for each separate GP practice code every three months.^6^ However, previous work has identified practices that operate through separate practice codes but which are in fact part of a single larger organisational entity.^2^ For some practices, this is explicit and transparent on practice websites, and will often contain the same practice name (e.g. Barclay). For others, it is not immediately obvious whether practices are operating as part of the same entity. Using data for July 2025, we identified practices that are associated with one another in this way using medical practitioner registration data. Specifically, we examined the unique General Medical Council (GMC) registration number of medical practitioners in Scotland in order to identify instances where a doctor was listed as a performer or salaried performer in more than one practice code. We then summed the practice list sizes for practice codes associated in this way. However, it is possible that this method overestimates the size of some megapractices if some doctors are performers or salaried performers in two otherwise completely separate practices. For one practice (Barclay) we decided to exclude some practice codes for this reason as we felt it was more likely that these were genuinely different organisation entities (not least because unlike some of the other megapractices, Barclay clearly links its practices together on shared websites).

This notwithstanding, it is more likely that this method will underestimate the size of some megapractices as there is evidence that some practices become part of the same structure without GPs from the megapractice being listed as performers across the practice codes. The Public Health Scotland database describing payments to General Practices provides notes on changes to practice codes, mergers between practices, and where practices have taken over others.^7^ This shows some additional practices being drawn into megapractices which are not captured by our method, but this does not provide a means of identifying all practices which operate as part of a megapractice.

### Interviews with key informants

We invited people listed as GP performers, and practice managers, in six megapractices to participate in interviews. Unfortunately, none responded and we were therefore unable to use those interviews to check our understanding of the practice codes that constituted megapractices, nor were we able to better understand their service model. We did complete five interviews with staff in non-megapractices, but as there was no comparison data from megapractices, we do not report the results of the interviews here. We have made four of these anonymised interviews available on the University of Glasgow’s Enlighten platform for other researchers to use (the recording failed on the other interview).

### Commercial profiles of GP megapractices in Scotland

We developed commercial profiles of a small number of practices in order to better understand the structure and business models, following methods developed to investigate commercial provision of healthcare services in other studies.^8^ We selected practices on the basis of the size (including several megapractices) and where markedly different models were drawn to our attention in interviews or by stakeholders: Barclay, Alba, Ayrshire Medical Group and OneMedical. The latter, which operates two practices in Scotland, was included on the basis that if its English operations were included it would be considered a megapractice. We collected data on the history and extent of each practice, the financial interests and arrangements, and the model of service delivery. Data was obtained from practice websites (including previous versions using specialist software); internet searches for relevant news articles; job and contract advertisements; social media posts; legal documentation; and Companies House records. For the latter, company records were cross-checked with practice names and GP performer names to identify all current and former registered companies, their financial records, all director names, and any details of the activities of associated companies.

### Creation of a typology of GP practices

The creation of a practice typology broadly followed the approach detailed by Stapley et al.^9^ Our data consisted of the PHS datasets, our limited interview data, and our commercial profiles. We used the profiles as our initial cases, detailing their salient features. We then tabulated these to determine areas of similarity and difference before refining our categories through discussion.

### Ethics

This project was approved by the University of Glasgow College of Medical, Veterinary & Life Sciences ethics committee (#200240040).

## Results

### Update of trends in the size of GP practices in Scotland

Using only individual practice codes, the median practice list size was 6,279 patients, and the average 6,822 patients, with the inter-quartile range being 3,783 to 8,948 patients (Figure 1). There were eight practice codes that were outliers,^a^ with only one code over 30,000 patients (at 64,558 patients). When associated practice codes were grouped, there were five groups (henceforth termed ‘megapractices’) with a total list size of over 30,000 patients (Figure 2). The largest by some margin was Barclay, with 119,810 patients across seven separate practice codes (having grown from 101,392 patients in April 2023^2^). Alba was next largest with 72,812 patients across five codes, then a group in Lothian (Blackhall, Murrayfield, Murieston and Liberton) with 52,577 patients across five practice codes, then Newburn with 45,458 patients over two codes.

**Figure 1:**
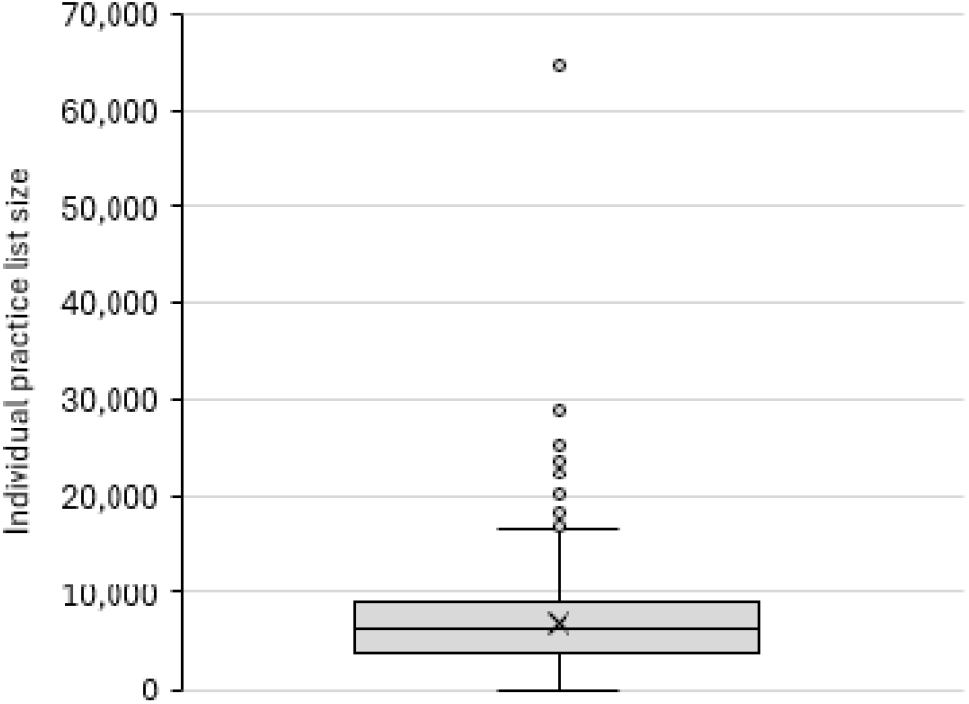
Distribution of practice list sizes across individual practice codes, Scotland, July 2025.

**Figure 2:**
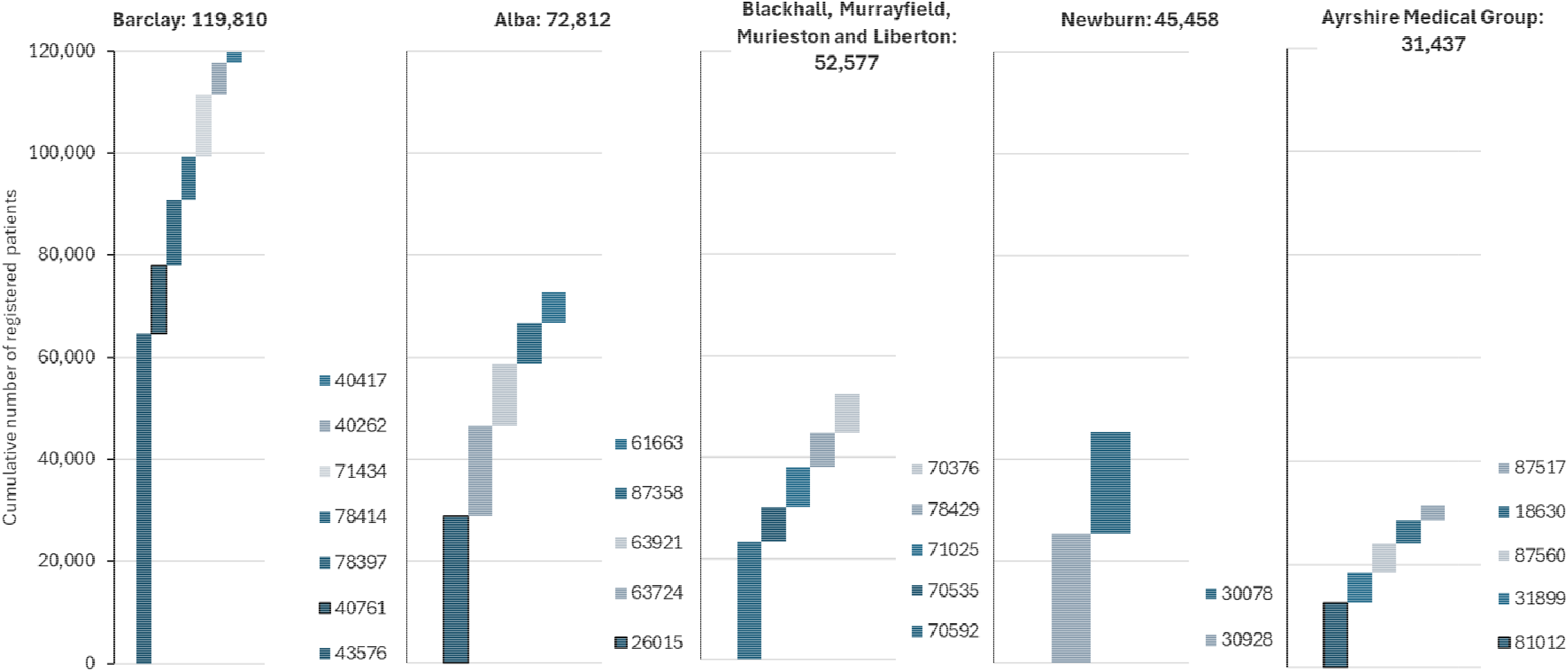
Breakdown of Scottish GP practice codes and cumulative list sizes for GMC-number associated practices with a total of >30,000 patients registered in July 2025*. * For the Barclay group, the following practice codes were not included as it seemed unlikely that the associated GMC numbers represented a genuine shared institutional entity: 30218, 40173, 43167 and 71434. For the Ayrshire Medical Group there are several additional practices which have been reported to be part of the same group which are not included here because they do not share any GPs with the same GMC number.^12^

Using our list size method alone, the Ayrshire Medical Group counted 31,437 patients across five codes, however, we are aware of a further six practice codes (Saltoun, Links, Valleyfield, Kinghorn, An Caorann, Central Buchan) that are part of the same group from news reports and job advertisements on gpjobs.nhs.scot^10,11^ which account for a further 31,437 patients, making a total of 63,501.

There were a larger number of individual or linked practice codes with total list sizes of 20-30,000 patients, including one covering patients in the Forth Valley area of 29,143 patients, and the Skene Medical Group with 28,630 patients.

### Commercial profiles

Our commercial profiling revealed a recent step-wise, and often rapid, accumulation of practices into large megapractice groups. Several megapractices spanned geographical locations, including multiple health board areas. For some megapractices there was evidence of complex financial arrangements for their leading GPs. However, the legal status, the number of practice codes that form part of the same organisational entity, financial arrangements and service model for several of the megapractices was not clear. There is currently no publicly available information which provides sufficient detail to confidently describe the emergent range of megapractices despite them providing NHS GP services for a growing proportion of the Scottish population. Some practices, such as Barclay, do not seem to be registered as Companies, and so more of the information available is from their self-descriptions on their websites and the writing of their leaders. Others, such as Alba, the Ayrshire Medical Group and OneMedical, have GP leaders with highly complex histories of multiple company ownership and dissolution, with a lack of clarity even within Company House records of the entities which provide GP services or hold NHS contracts. Ascertaining the actual list size of some megapractices was also difficult (e.g. only around half of the total list size was identified through common GMC numbers for the Ayrshire Medical Group, with the other practice codes only identified through news reports and job advertisements, suggesting that substantial undercounting for other practices is also possible). We also identified evidence that several of the GP leaders within the megapractices have companies which appear to be in conflict with public health goals (e.g. ownership of fast-food companies).

**Table 1:** High-level summary of commercial profiles.

| **Megapractice** | **History and growth** | **Ownership and financial interests** | **Service model** |
| --- | --- | --- | --- |
| Barclay | Barclay started in two sites in Glasgow before their rapid expansion, justified by the group in a BMJ comment as a means of preventing ‘collapse’ of GP provisioning in some areas.<sup>4</sup> Three practice codes in West Lothian came under the Barclay group in 2017,<sup>6</sup> and there was amalgamation of Glasgow practices into a smaller number of codes by 2023.<sup>6</sup> As of July 2025, there are six practice codes bearing the Barclay name over nine sites (Clydebank, Maryhill, East Craigs, Carmondean, Fauldhouse, City, University, Maybury and Harbourside (which has three sub-branches: Clydeside, Victoria Park and Kingsway)).<sup>5</sup> It is the largest practice in Scotland with almost 120,000 patients in July 2025. | As Barclay does not appear to be registered as a company, there is little information available on its financial arrangements. One practice partner has commented to media on an aspiration for Barclay to, “...become like a social enterprise, a bit like John Lewis”, with capped earnings for GP partners and profit shares for all staff.<sup>3,13</sup> | We characterise as GP-light, with multiple AHPs employed to undertake house calls, provide telephone and face-to-face appointments, and condition-specific support. GP resources are centralised and used for triage and the most complex and serious cases. It is unclear which staff work for Barclay generally, or in each practice specifically, and the extent to which continuity of care is possible is unclear. New patients are not prescribed a range of drugs deemed by the practice as ‘red drugs’.<sup>b</sup> Provides specific care for patients not registered in the practice (e.g. long-acting contraceptives) which attract additional payments. |
| Alba | Until 2017, this was a small practice in Lanarkshire. In 2018 the practice started to grow quickly, with six other practices being absorbed.<sup>2</sup> By 2025, Alba self-describe three groups of practices centred in | No companies are registered with the name Alba or Lanarkshire Medical Group, but the practices name two GP performers)<sup>6</sup> who are connected to a large number of private limited | We characterise as GP-light, with multiple AHPs employed to provide services for its patients beyond GPs. There are only two GP performers in Alba, suggesting a high number of |

<sup>b</sup> Listed as: tramadol, dihydrocodeine, pregabalin, oxycodone, 30/500 co-codamol, gabapentin, benzodiazepines, and ‘z-class’ sleeping pills.<sup>14</sup>

|  |  |  |  |
| --- | --- | --- | --- |
|  | Lanarkshire (Strathaven, Alison Lea, Greenhills, Woodstock and Coatbank), Forth Valley (Hallpark, Bannockburn, Kersiebank and Westburn) and Glasgow and Clyde (Bargarran and Quarrieside), covering 11 practices in total across a common website.<sup>15</sup> The Lanarkshire practices were sometimes described as the 'Lanarkshire Medical Group' (LMG), and the Forth Valley practices as the 'Forth Medical Group' (FMG).<sup>16</sup> | companies, both current (n=14) and dissolved (n=10). Several of these companies could plausibly be trading firms for providing GP services (e.g. Meducating Ltd. and ADM Consultancy Ltd.) and therefore be the financial entity under which the GP services are being run. In addition, both doctors have had financial interests in dispensing pharmacies (most of which they have withdrawn from), a fast-food company (Chillicious Ltd.) and several property firms. | salaried and locum GPs. One of the practices (Woodstock) was investigated after patients complained about being unable to make appointments.<sup>17</sup> On the common website for all practices, specific doctors are identified in each practice location, although with no information on their qualifications, interests or working patterns. |
| Ayrshire Medical Group (AMG) | In October 2023, AMG was reported to have taken over six additional practices.<sup>10,11</sup> but none of the GPs working in any of the pre-existing AMG practices were listed as working in any of these new practices in July 2025,<sup>6</sup> raising the prospect that other practices may be part of AMG but are not identifiable through common GMC numbers. | One GP is listed as a performer in five of the practices and owns a 10% stake in AMG Scot Ltd., but it is unclear what assets this owns nor what services it runs. The other 90% stake in AMG Scot Ltd. is owned by a non-GP but who shares the same surname and who is additionally sole owner of Zaelno Meded Ltd. and Zaelno Ltd., although the nature of these firms is unclear. In addition to the work with AMG, the listed GP reports in online biographies that they undertake work via 'Medicspot' (a<br>private online weight loss service), although it is unclear how current this is. | There is limited evidence to clarify the service model but there seems to be high use of locum GPs to provide services in several locations. |
| OneMedical | OneMedical (also known as OneMedicare) run two practices in Aberdeen (Great Western and Whinhill) as a Scottish arm of the much larger OneMedical group, with just under 20,000 patients in total. Its wider portfolio includes two urgent care centres, a community dermatology service, at least ten GP surgeries, and two walk-in centres, mainly in England. It describes itself as a “nationwide and family-owned health group, providing NHS primary and urgent care services, as well as designing, building and managing medical premises”. | The group includes an array of companies (including OneMedical Group Scotland and a property firm), and ownership differs slightly across the different companies, but a family name is common across firms. | There is little detail about the service model available, but salaried GPs rather than GP partners seem to provide most of the medical input, and no details of the interests or qualifications of the GPs is provided on the practice websites. |

### A tentative typology of GP practices in Scotland

From the available data, we situate our updated understanding of megapractices in Scotland within a contemporary, but tentative, typology of GP provisioning. The key aspects of commonality and difference across practice types were identified in relation to: ownership, legal status and motives; the extent of service provision by doctors or AHPs; and practice list size.

#### Ownership, legal status and motives

Until relatively recently, almost all GP practices in Scotland operated as voluntary partnerships whereby small groups of GPs would form partnerships and contract with the NHS. Few, if any, were constituted as limited companies. The use of multinational companies to provide GP services as has been seen in England for many years^18^ was stalled in Scotland after a failed bid by Serco to run a practice in Lanarkshire^19^ and a subsequent change in legislation.^20^ Yet, the limits of that legislation have seen the recent emergence of several large megapractices which operate as limited companies, as well as some smaller practices (e.g. https://deucharwilliamshealth.com/), and at least one larger company opening a Scottish arm (OneMedical). In the other direction, there are now examples of more socialised provisioning models, including directly run NHS practices (under the 2C contract), a GP workers’ co-operative (Newfield practice in Dundee) and a Community Interest Company (CIC, Riverdene practice in Aberdeen). It is notable that workers’ co-operative and CIC models do not necessarily preclude large salaries, but are generally understood as structures which prevent profiteering and which foreground social objectives. We have also identified evidence of several GPs having multiple financial interests including pharmacies, property (residential and practice premises), commercialised healthcare or wellbeing advice provision, and fast-food.

#### Service provision

The 2018 General Medical Services contract and Primary Care Improvement Fund led to increased multi-disciplinary working in primary care with pharmacists, physiotherapists and other AHPs working in practices, but employed by Health and Social Care Partnerships.^21^ However, some practices also directly employ a wide range of AHPs, including advance nurse practitioners, physiotherapists, mental health nurses and paramedics. In this case, the model can often be described as ‘GP-light’, whereby patients are steered towards AHPs and the GPs see only the most serious or complex cases, with more triage and remote consultation than under the traditional model. In the GP-light model, salaried GPs and locum GPs seems to be used more heavily than in the traditional model where most of the medical staff are GP partners. Greater understanding of the variation in the GP-patient ratio, and the form of GP (partner, salaried, trainee, locum), is needed. With the GP-light model, it is not clear whether continuity of care is possible, with often very large numbers of patients managed across multiple sites by remote GPs. Some megapractices have seen repeated patient complaints about a lack of access to GPs, although this has been difficult to disentangle at times from whether this is due to the GP-light model, or the consequences of pre-existing GP vacancies which led to the contract being awarded to the megapractice. Megapractices are also generally characterised by operating across multiple locations (beyond a simple branch practice in a neighbouring area), practice codes and health boards. Some practices (e.g. Barclay) have implemented restrictions on new patients joining in relation to the drugs that they will prescribe which might disincentivise particular patient groups and create selective recruitment pressures. Barclay also has created additional service offers to people not on their practice lists for some services (such as long-acting contraception). The implications of new models of provisioning for training (including medical students and GP trainees) are unclear.

#### Practice list size

Although most practices remain of modest size (with half of individual practice codes having an associated list size of between 3,783 and 8,948), there has been the emergence of some very large practices (not least Barclay and Alba) driven by two trends. First, NHS Boards have experienced difficulties in finding partners to take over practices as GPs retire (and GPs approaching retirement have struggled to recruit partners to achieve continuity). Second, some practices have chosen to amalgamate to provide greater resilience and scale, although we have identified less evidence to describe this trend clearly.

#### A tentative typology

Using these three key dimensions, we propose a typology comprising four models: traditional practices (17j and 17C), social enterprise practices, direct NHS provision (2C practices) and megapractices. The vast majority of Scottish practices fit the traditional type most closely, although this is also the type which is declining most rapidly. There are now at least two examples of social enterprise practices in Scotland, with one workers’ co-operative and one community interest company. These are distinct from traditional practices because of their legal status and clear rejection of the private enterprise contractor model,, but are otherwise quite similar. Direct NHS provision is also distinct because the staff here are salaried and directly employed by the NHS, usually on the understanding that this is an interim step taken by NHS Boards until services can be contracted out again. However, there are also examples where this form of provisioning is used deliberately to meet a particular need, in line with existing theories of direct public provisioning.^22^

Megapractices—large and rapidly growing multi-site practices that tend towards a GP-lite model of provision—are becoming prominent fixtures in Scottish primary care but nonetheless display substantial variation in ownership structures and service model. For example, there is marked variation whether they utilise company legal status for one or more entities associated with the GP partners, the financial rewards for the owners and GPs, whether there are restrictions for prescribing for new patients, and the extent to which additional services are provided (including clinical services and other financial interests such as property). There are also some examples of small practices which have adopted company legal status which do not fit clearly into any of our proposed types. It may be in due course that further examples emerge which allow further disaggregation of the megapractice category.

**Table 2:** A tentative typology of GP practices in Scotland.

| **Practice type** | **Ownership, legal status and motives** | **Service model** | **Practice list size** | **Examples** |
| --- | --- | --- | --- | --- |
| Traditional | Voluntary partnership, earnings in line with medical pay in NHS. | GP-led with incorporation of some AHP provision. Continuity of care prioritised. (17c and 17j practices) | Mostly under 10,000 patients. | Majority of Scottish practices |
| Megapractice | Mostly registered as companies, but some important exceptions (e.g. Barclay). | GP-light with extensive use of AHPs. Operates across multiple sites and health boards. Continuity of care unclear. Variation in: profit sharing/earnings; ability to access appointments; prescribing restrictions for new patients; and additional service provision. | Usually over 20,000 patients, but some over 100,000. | Barclay, Alba, Ayrshire Medical Group, OneMedical |
| Social enterprise | Clear social motivation with absence of financial motivation for owners. | GP-led with incorporation of some AHP provision. Continuity of care prioritised. | Mostly under 10,000 patients. | Newfield (workers' co-operative) and Riverdene (community interest company) |
| Direct NHS provision | Practice owned and run by NHS with staff employed directly. | GP-led with incorporation of some AHP provision. Continuity of care prioritised. (2C practices) | Mostly under 10,000 patients. Some specialised practices provisioning for specific groups (e.g. nursing homes or homeless). | Edinburgh Access Practice |

## Discussion

GP megapractices have emerged in Scotland without overt debate nor planning. The largest megapractices now provides for around 120,000 patients, with four others providing for over 30,000 patients. Several of these megapractices operate as companies and with complex legal structures and multiple financial interests for their GP directors. However, we struggled to engage GP from megapractices in our research, and so there remains substantial uncertainty about their true extent, legal arrangements, financial interests and service models. We tentatively propose a typology for Scottish GP practices on the basis of the available data to include: traditional practices, megapractices, social enterprise practices and direct NHS provision, recognising that there is substantial diversity and uncertainty within the megapractice type, and there are some practices which do not fit easily into any category. The lack of transparency about the emerging practice models, and the lack of evaluation of their impacts on service access and health outcomes, should urgently be addressed.

Our approach is based on the best available data published by Public Health Scotland, and attempts to engage with practices directly to understand them. However, there are a number of important limitations we encountered. First, it is recognised that there are approximately 10% more people registered in Scottish GP practices than the estimated population size, which is likely to be partially due to people not de-registering with practices when they move outside of Scotland.^23^ Second, we recognise that we may have incorrectly grouped practices together as megapractices where GPs work in two separate practices which are entirely independent. Where we have identified this risk (e.g. through examination of practice websites) we have kept those practices separate in our analysis. More likely is that some practices which are actually part of a megapractice have not been coded as such because there is no common GP listed across practice codes, thereby underestimating the scale of some megapractices. Third, GPs and practice managers working in megapractices did not engage with our requests for interviews, and so may have inadvertently misrepresented the characteristics of some practices. Fourth, our suggested megapractice type contains substantial variation, but we were unable to sensibly disaggregate this because of a limited range of examples to otherwise group together.

Similar to the trends we describe here for Scotland, there has been a rise in megapractices in England, with 7% of practices there being incorporated companies, and 2.3 million patients listed in practices with over 100,000 patients, by 2023.^24^ Very little is known about the impacts of moving towards megapractices,^25^ or large-scale collaborations between general practices.^26^ There is some evidence that larger practices do better on some markers of care quality, but this seems to be explained by much greater variation for very small practices (in particular single-handed practices).^27,28^ Globally, there is evidence that privatisation of healthcare leads to selective patient intake, reductions in staffing, and worse health outcomes.^29^

Furthermore, greater marketisation in healthcare provision has long been theorised to exacerbate access inequalities (i.e. the inverse care law).^30^ The extent to which the practice types identified here, in particular megapractices, reflect the forms of privatisation and marketisation that might lead to these consequences is unclear, although the commercialisation of primary care across the UK has been underway for some time, facilitated by changes in contracts which date back to 2003.^18^ Although large multinationals and companies have operated in England for some time, this has until very recently not been the case in Scotland, not least because of the Tobacco and Primary Medical Services (Scotland) Act 2010 which dictated that at least one member of the contracted entity had to be a medical practitioner.^20^ It seems that the intention of this Act to reduce commercialisation of general practice in Scotland from outwith is increasingly being circumvented by GPs setting up companies and merging practices together (i.e. commercialisation from within). A new GP contract was announced at the end of 2025 with additional funding for practices,^31^ but the implications for GP practice types and GP recruitment remain unclear.

Scottish policymakers should carefully consider the risks of allowing current trends towards megapractices without a deep understanding of the financial and legal structures of these practices, their service models, and the implications for undergraduate and postgraduate medical training, healthcare access and continuity of care,^32^ health outcomes and inequalities. The potential for conflicts of interest to arise, including the potential incentives to minimise costs at the expense of provision in order to inflate salaries or company dividends, and the ownership of other businesses which have the potential to conflict with the interests of the NHS or patients (e.g. ownership of a fast-food company, ownership of community pharmacies, healthcare property ownership, private medicine provision) is real and should be carefully considered. Data systems should be improved so that common ownership is transparent across practices and health boards. Further research to understand the impacts of the new models of GP provisioning on healthcare access, health outcomes and inequalities is urgently needed.^2^

## Conclusion

The shape of GP practice provision in Scotland is continuing to change, with a trend towards megapractices. Several megapractices operate as companies with complex legal structures and new risks of conflicts of interest. GP-light models of service delivery are becoming more common without sufficient understanding of the impacts on healthcare access, health outcomes or inequalities. Policymakers should carefully consider: the risks of the current drift in approach to provisioning; commissioning research to evaluate the impacts of new models; mandate greater transparency in legal and financial arrangements amongst entities holding NHS contracts; and should be prepared to legislate to ensure provisioning meets public needs and interests.

## Data Availability

We have made four of these anonymised interviews available on the University of Glasgow Enlighten platform for other researchers to use (the recording failed on the other interview).

https://eprints.gla.ac.uk/

## Acknowledgements

We would like to express our sincere thanks to the GP staff who generously gave their time to complete interviews for this paper.

## Footnotes

a Outliers were defined in the standard way, as points lying above (or below) the upper (or lower) quartile by 1.5 times the inter-quartile range. In this instance this meant that there were no outliers of small practices because the lower value crossed zero.

